# A population-based resource for the prioritization of rare variants of unknown significance in clinical genomics

**DOI:** 10.64898/2026.11.03.26367175

**Authors:** Sarah Tremblay, Claudia Moreau, Gilles-Philippe Morin, Catherine Laprise, Simon L. Girard

## Abstract

Rare diseases represent a major public health challenge because they are often associated with severe phenotypes, yet many causal variants remain undiscovered. Founder populations provide a unique opportunity to identify disease-causing variants through the local enrichment of rare alleles. Using whole-genome sequencing data from individuals in the Saguenay-Lac-Saint-Jean population, enriched variants were identified and their founder status was assessed through identity-by-descent sharing. Variant impact was evaluated using functional prediction tools without relying on prior clinical classification. A total of 131 founder variants of interest were identified, most of which would likely have been overlooked by conventional approaches based on existing clinical annotations. We developed a web-based platform that facilitates clinician access to the latest interpretation of their genetic variants of interest. These findings demonstrate that potentially pathogenic variants can remain undetected even in extensively studied founder populations. More broadly, they highlight the value of population structure-based approaches for uncovering rare disease variants and improving the characterization of genetic disease risk across human populations.

## Introduction

Rare diseases tend to have more severe clinical consequences than common conditions[1]. Although each disorder affects only a small number of individuals, their combined prevalence is substantial, making rare diseases a significant public-health concern and a considerable burden for patients and families[2, 3].

The major challenge of studying rare disease-causing variants lies in their low frequency in the broad population, which makes them difficult to detect and characterize. Founder populations have deeply contributed to overcome this obstacle: their demographic history increases the frequency of certain rare variants[4], creating a context in which both these variants and their associated phenotypes become observable[5–8].

Most rare disease studies rely on clinically well-defined variants catalogued in databases such as ClinVar, for which the associated phenotypes are already established[9]. However, focusing primarily on such variants limits the identification of novel pathogenic variants, especially those lacking clinical evidence for classification and therefore categorized as variants of uncertain significance (VUS). Consequently, variants contributing to atypical or unrecognized phenotypes may remain overlooked. This challenge is reflected in clinical practice, where approximately 22.5% of exome and genome sequencing results remain inconclusive because of VUS[10].

These limitations highlight the need for population contexts where rare pathogenic variants can be detected independently of prior clinical classification. Saguenay–Lac-Saint-Jean (SLSJ) region in Quebec, Canada, represents one of the most thoroughly studied founder populations[11, 12]. Despite decades of genetic research and the identification of numerous disease-associated founder variants, the clinically relevant variation present in this population remains incompletely characterized[13] and recent studies continue to uncover enriched pathogenic founder variants in the SLSJ population[13–16], suggesting that additional clinically relevant variation may remain undetected.

This study aims to identify novel founder variants in SLSJ by leveraging population structure rather than clinical classification, and to evaluate their potential functional impact and clinical relevance. In founder populations, patterns of IBD sharing can reveal variants introduced by a common ancestor, as these variants typically reside on shared haplotypes. Combined with evidence of population enrichment, these signatures provide a powerful means of detecting rare variants independently of existing clinical annotations or classification. To support the translation of these findings into clinical practice, this project will also develop a web-based resource enabling clinicians to rapidly access the population and genetic context of encountered variants, thereby improving the prioritization and interpretation of variants with potential clinical significance.

### Data and methods

Detailed methods can be found in Supplementary material.

## Results

Potential founder variants in the SLSJ were identified in whole-genome sequences (WGS) from 817 individuals from the SLSJ and 1,696 from elsewhere in Quebec according to multiple selection criteria (See methods). Based solely on their increased frequency in individuals from the SLSJ region compared with other regions of Quebec (≥10%), more than 11 million variants were initially identified as potential founder variants (Fig. 1), highlighting the need for additional criteria. Among these, only 0,4% were both located within the Consensus Coding Sequence (CCDS) and carried by enough individuals for downstream analyses. Following all founder variant criteria, 14,621 variants remained, underscoring the need for additional prioritization criteria. Of these, 5,96% were predicted to be likely pathogenic by AlphaMissense, while 0,896% had additionally a high pLI score. To further prioritize variants with a higher likelihood of functional impact, we focused on variants located in genes intolerant to loss-of-function mutations (pLI >0.9), yielding a final set of 131 candidate variants (Fig. 2; Supplementary Table 1).

**Fig 1.**
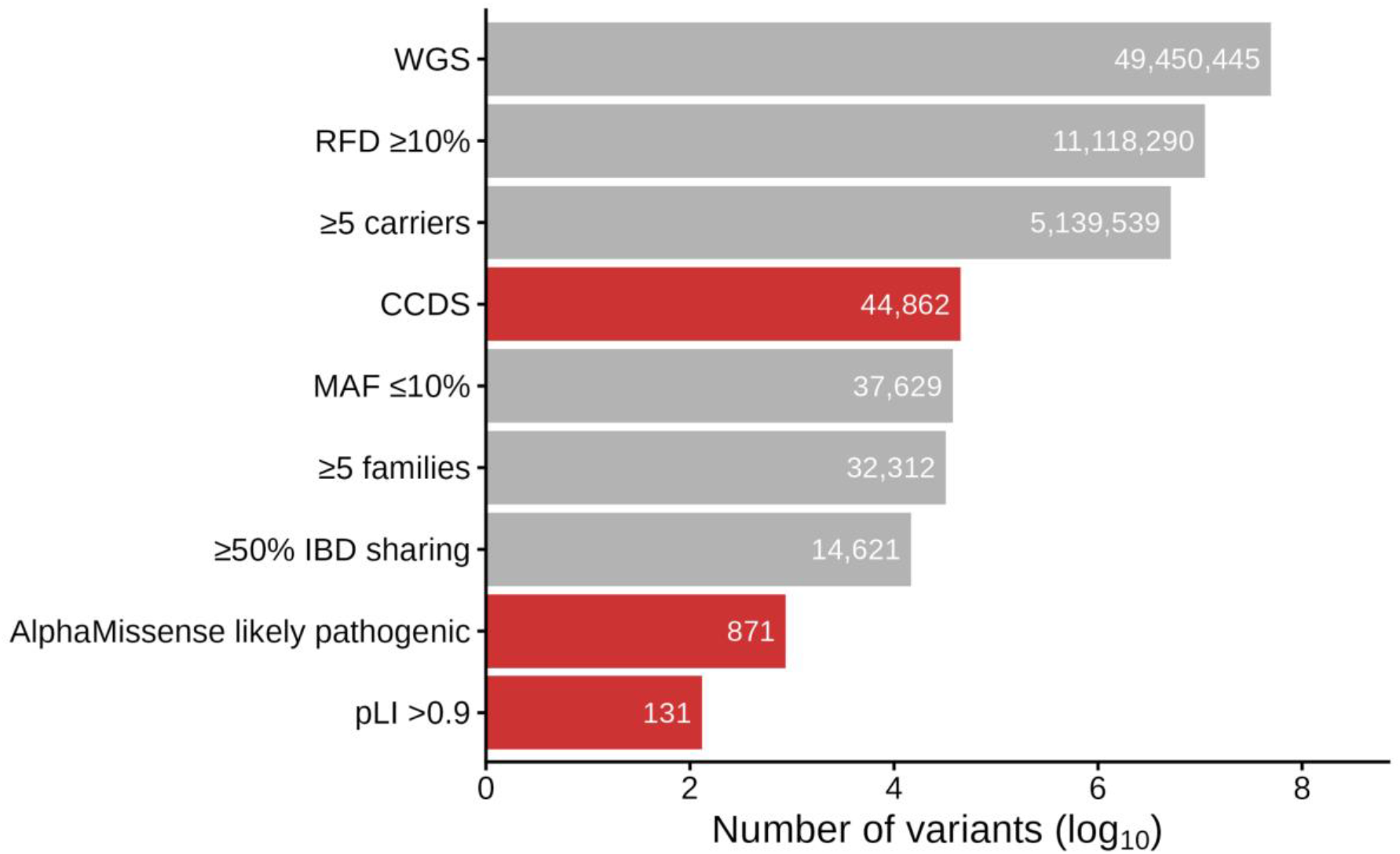
Number of variants remaining after each filtering criterion. Grey criteria are related to founder status, whereas red criteria corresponds to filters associated with the likelihood of modifying the protein.

**Fig 2.**
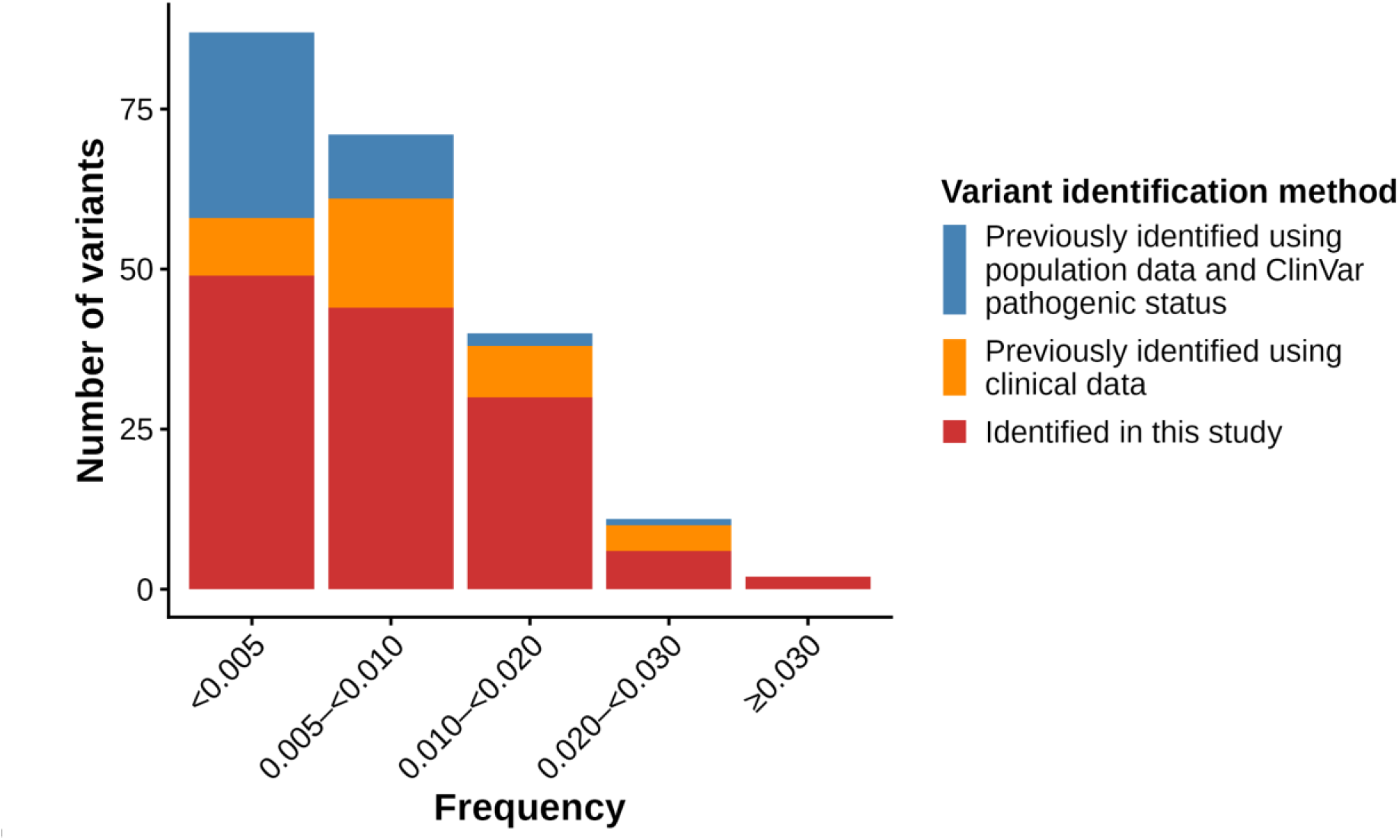
Distribution of founder variants by frequency bin and identification approach.

The frequencies of the 131 founder variants predicted to modify the protein identified in this study were compared with those of founder variants previously discovered through population-based studies, clinical data or ClinVar pathogenic classification[6, 11–13, 17] (Fig. 2).

## Discussion

The approach used in this study enabled the identification of numerous previously undocumented variants of potential interest in SLSJ. These findings demonstrate that leveraging population structure rather than clinical classification uncovers variants that would otherwise remain undetected due to mild phenotypes or low frequency in the broad population, which limits their inclusion in clinical databases[9, 18].

More broadly, the approach uncovered 131 founder variants with potential protein-modifying effects. Remarkably, despite their higher frequency in SLSJ, these variants had remained undetected even in this thoroughly studied founder population[11, 12]. This represents a substantial number when compared to the 72 previously known disease-associated rare variants[6, 11, 12, 17], or even the 42 reported in a recent population-based study[13].

As these newly identified founder variants have frequencies similar to those of disease-associated variants, it is plausible that the burden of rare diseases in SLSJ may be greater than previously recognized[6, 11–13, 17]. However, many of these variants would likely remain unnoticed under traditional rare variant identification approaches. Rare variant studies typically prioritize variants classified as pathogenic or likely pathogenic in ClinVar[5, 13]. Under such an approach, the variants identified here would have been overlooked despite their founder status, as most are absent from the database or classified as VUS. Although some are currently classified as benign or likely benign in ClinVar, their predicted impact on the protein remains noteworthy and justifies keeping them under consideration, particularly given that variant classifications may be revised as new evidence emerges[19].

To facilitate variant’s interpretation as most are absent from ClinVar, a web-based resource was developed as part of this project, providing clinicians with direct access to population-based metrics and the likelihood of the variant to be pathogenic. This resource represents an important step toward addressing the challenge posed by VUS, which continues to be a major obstacle in clinical practice. By facilitating the identification of recurrent variants across clinical reports, the resource may support variant interpretation and help prioritize candidate variants for future functional studies.

Some limitations should be considered when interpreting these findings. First, this approach is well suited for detecting recessive founder variants, but has limited ability to identify dominant variants. This limitation is illustrated by myotonic dystrophy type 1 (DM1), a dominantly inherited genetic disorder enriched in SLSJ, yet present in only 0,16% of participants in CaG[20]. Given the minimum carrier threshold applied in this study, it is therefore plausible that unidentified dominant disease-causing variants would occur at frequencies too low to be retained for analysis. Second, the lack of functional validation limits the interpretation and classification of the identified variants, as no phenotypic evidence is available to support a causal relationship between a variant and disease.

Despite these limitations, the present study provides a more comprehensive portrait of the genetic variation present in the SLSJ population. By reporting and documenting numerous previously unreported rare founder variants in a web-based resource, this study offers a valuable tool for future research aimed at assessing their clinical significance and potential association with rare diseases. Some of the currently unexplained phenotypes observed in the clinics may be attributable to these variants, highlighting the importance of documenting them. More broadly, these findings demonstrate that substantial genetic variation can remain unreported even in a thoroughly studied founder population such as SLSJ. This suggests that similar hidden variants may exist in other founder populations for which the presented approach could be applied.

## Data Availability

Genotyping, WGS and imputations of CaG participants are available under restricted access from CaG biobank due to the informed consent given by study participants (https://cartagene.qc.ca/en/researchers/access-request.html). Data from the SLSJ asthma family cohort are available in the Canadian Dataverse Repository Borealis on reasonable request (https://doi.org/10.5683/SP3/QEDOPE). Genealogies from BALSAC population register used for classification are available upon request at https://www.balsac.ca/, following the Policy on Access to BALSAC Data for Research Purposes.

## Web resources

The web resource is accessible at https://genopop.github.io/Founder_variants_likely_to_modify_protein/

## Data availability

Genotyping, WGS and imputations of CaG participants are available under restricted access from CaG biobank due to the informed consent given by study participants (https://cartagene.qc.ca/en/researchers/access-request.html). Data from the SLSJ asthma family cohort are available in the Canadian Dataverse Repository Borealis on reasonable request (https://doi.org/10.5683/SP3/QEDOPE). Genealogies from BALSAC population register used for classification are available upon requestat https://www.balsac.ca/, following the Policy on Access to BALSAC Data for Research Purposes.

## Code availability

The codes used for this study are available in the following GitHub repository: https://github.com/Genopop/Founder_variants_likely_to_modify_protein

## Acknowledgements

We are extremely grateful to all participants in this research. This work was supported by the Digital Research Alliance of Canada which provided access to storage and computing resources. Mylène Gagnon curated the WGS and imputed datasets, while Anne-Marie Madore proceeded curated genotype data and performed DRAGEN processing of the WGS data.

## Author contributions

ST drafted the manuscript. All authors approved the final version of the manuscript. ST and CM conducted the analyses and interpreted the results. GPM contributed to the calculation of IBD sharing. CM, GPM CL and SLG revised the manuscript.

## Ethical approval

This study was approved by the Research Ethics Committee (CER) of the Centre Intégré Universitaire de Santé et de Services Sociaux (CIUSSS).

