## Supplemental material for "A population-based resource for the prioritization of rare variants of unknown significance in clinical genomics"

**Supplementary data and methods**

Cohorts

The CARTaGENE (CaG) and Saguenay–Lac-Saint-Jean (SLSJ) asthma family cohorts were used in this study[1–3]. The CaG cohort is composed of 43,031 participants aged 40 to 69 and recruited randomly from 2009 to 2011 and from 2012 to 2015 in the Quebec province. The recruitment took place in six metropolitan areas: Montreal, Quebec, Trois-Rivières, Gatineau, Sherbrooke and Saguenay. The cohort includes 2,173 whole genome sequences (WGS) and 29,337 genotypes. Self-reported phenotypes for this cohort are also available, including different health statements such as disease occurrence. The SLSJ asthma family cohort includes 1,394 individuals recruited from 1997 to 2007 in SLSJ, coming from 271 families, of whom 1,214 were genotyped.

Data cleaning and imputation

Production of WGS from the CaG cohort is described in the CaG documentation[4]. WGS from the SLSJ asthma family cohort were produced at Centre d'expertise et de services Génome Québec using the DRAGEN Germline pipeline, with reads aligned to the hg38 reference genome and variants called using the DRAGEN Small Variant Caller. Using GVCF files of both cohorts, a joint variant calling was performed using DRAGEN PopGen Joint Genotyping workflow via the Population explorer WebApp V4.0, producing a merged VCF file. These joint called WGS were cleaned using PLINK version 2.0[5]. After splitting biallelic and multiallelic variants, those with more than 5% missing genotypes (--geno 0.05) were filtered out. Mendelian errors were set to missing using --set-me-missing.

NON_REF and monomorphic variants were removed. Biallelic and multiallelic variants were then merged. Remaining variants in centromeric regions were removed, as well as those in ENCODE Blacklist regions. The final VCF file contains 49,450,445 autosomal positions and 2,588 individuals, of whom 699 belong to the SLSJ asthma family cohort and 1,889 to the CaG cohort.

Genotyping data cleaning of the CaG cohort is described in Michel et al.[6]. Genotyping data from the SLSJ asthma family cohort were cleaned using PLINK[3, 5]. 304 individuals with more than 5% missing SNPs were removed, along with 9,435 SNPs with more than 5% missing genotypes and 221 SNPs with HWE p<1e-6. The final SLSJ familial dataset contains 1,197 individuals and 572,237 variants.

Imputation was performed separately for each genotyping batch using the joint WGS call set from both cohorts as the reference panel. Phasing of genotypes and WGS was performed using SHAPEIT5 following authors' suggested methodology[7]. IMPUTE5 v1.1.5 was used for imputation with a local reference panel to enhance imputation accuracy of rare variants[7–9].

Regional classification

To identify founder variants in SLSJ, individuals from this region were distinguished using a classification method, as recruitment location alone does not provide sufficient resolution to accurately define the population and reliably detect enriched variants[10].

Because supervised classification requires reference labels, we relied on the BALSAC population register, which links Quebec civil records to reconstruct genealogies and provide reliable ancestry profiles for registered individuals[11].

By leveraging individuals for whom both genomic and genealogical data were available, we trained a model to predict regional ancestry profiles from patterns of genetic relatedness calculated using total identity-by-descent (IBD, see next section) sharing between individuals[12]. The model was then used to infer ancestry profiles for participants lacking genealogical records, enabling the identification of individuals with ancestry patterns consistent with the SLSJ founder population.

The final SLSJ cluster includes 3,456 genotyped individuals, of whom 817 have a whole genome sequence.

#### Identity-by-descent sharing

Considering that a founder variant usually originates from a single ancestor in a population with a founder effect, and within a relatively recent time frame to increase in frequency due to drift, this variant will still be in LD with other surrounding variants and carriers will share not only the variant, but also the surrounding haplotype. Segments shared identical-by-descent (IBD) were used for this purpose.

Because the overlap across the different genotyping arrays resulted in a limited number of shared SNPs, we reconstructed a synthetic genotyping array from the imputed data. We selected 475,892 SNPs among the most frequently represented markers on the Global Screening Array (GSA), the platform used for most individuals. Genotypes at these markers were generated for all individuals from both CaG and SLSJ familial cohorts. Genotypes inconsistent with the corresponding true genotype data were excluded. Standard quality control procedures were then applied, including the removal of SNPs with >2% missingness or significant deviation from HWE (p<1e-6), as well as individuals with >2% missing genotypes. The final array includes 470,037 for 30,543 individuals. This synthetic array was phased using Beagle version 18May20 and identity-by-descent (IBD) segments were assessed using Refined IBD version 17Jan20[13, 14]. We retained only IBD segments of 2Mb or longer and with a LOD score greater than 3.

Selection of variants

Variants in the SLSJ cluster were identified in the WGS data according to multiple criteria based on either the founder status or a potential effect on the protein.

To assess the founder status, variants carried by at least five individuals were retained. To avoid selecting variants enriched in a limited number of families, each variant was required to be observed in at least five distinct families, supporting its distribution throughout the SLSJ population.  Variants with a minor allele frequency (MAF) ≤10% and having a relative frequency difference (RFD) ≥10%[6] between SLSJ and the rest of Quebec ), were selected. Finally, to verify that the identified variants originated from a single introduction into the population and were consistent with a founder event, we investigated the extent of IBD sharing around each variant, as described in[6]. Given the presence of families in the dataset, carrier pairs often shared an IBD segment on both of their haplotypes at the same location. To ensure an accurate estimation of IBD sharing linked to the variant of interest, the calculation was restricted to the haplotype shared by most of the variants' carriers. An IBD-sharing threshold of ≥50% at the variant locus was applied, following methods in [6, 15].

To enrich for protein-altering variation, we retained variants located in Consensus Coding Sequence (CCDS) regions and their 10 bp flanking intervals[16]. AlphaMissense was then used to flag variants predicted to have a deleterious effect on the encoded protein[17, 18]. These predictions do not correspond to clinical pathogenicity classifications such as those provided by ClinVar, but rather provide an estimate of potential functional effects. Finally, for variants predicted as likely pathogenic by AlphaMissense, gene intolerance to loss-of-function mutations was assessed using the pLI score. Only variants located in genes with a pLI score >0.9 were retained[19].

After identifying variants enriched by the founder effect and predicted to alter the encoded protein, phenotypes from the CaG cohort were examined to prioritize those most likely to have functional consequences. Because these variants have not been previously documented, their mode of inheritance cannot be inferred. Phenotypic assessment therefore centered on homozygous individuals, since we lack power to detect dominant variants as they would be much less frequent in the population. Although all available phenotypic information was reviewed, particular attention was given to severe or atypical manifestations, including surgical interventions, diseases and infections. Relevant phenotypes were evaluated considering the gene's function to determine whether a biological link would be plausible.

### Supplementary results

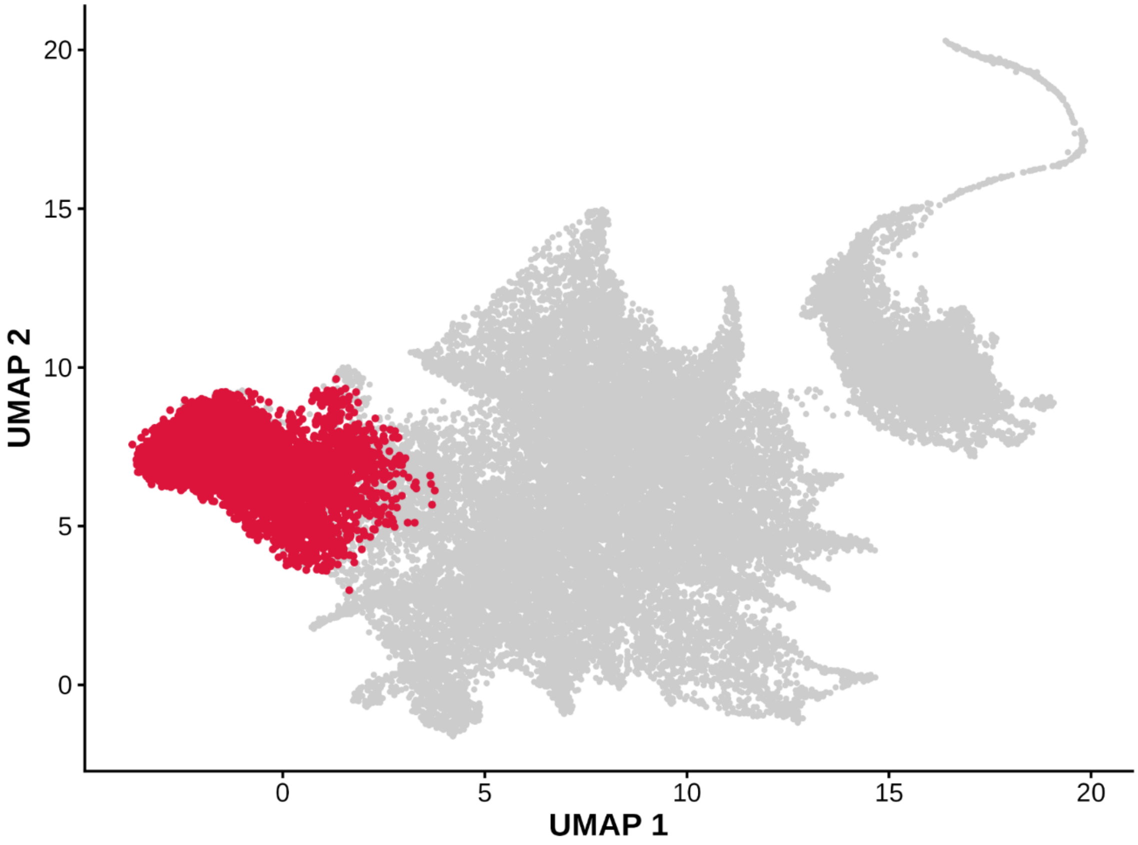


**Supplementary Fig. 1 UMAP of genetic coordinates of individuals from CARTaGENE and SLSJ asthma family cohort.** Two-dimensional UMAP based on the first 20 genetic coordinates of individuals' total IBD sharing and initialized using the first two PCoA. Individuals belonging to the SLSJ cluster are colored in red.

**Supplementary Table S1. Additional information on founder variants likely to modify the encoded protein**

See Excel file Tableau_131.xlsx
